# Development of a Decision Support Tool for Acute Appendicitis

**DOI:** 10.1101/2021.11.08.21266077

**Authors:** Joshua E. Rosen, Joshua M. Liao, David R. Flum, Jonathan E. Kohler, Giana H. Davidson

**Affiliations:** Surgical Outcomes Research Center, University of Washington, Seattle, Washington; Department of Surgery, University of Washington, Seattle, Washington; Decision Science Group, University of Washington, Seattle, Washington; Department of Medicine, University of Washington, Seattle, Washington; Division of Pediatric General, Thoracic and Fetal Surgery, Department of Surgery, University of California – Davis, Sacramento, California; Department of Health Systems and Population Health, University of Washington, Seattle, Washington

## Abstract

**Background:** Multiple randomized controlled trials have shown that it is safe and effective to treat appendicitis with antibiotics or surgery. There are no tools available to assist surgeons and their patients in choosing the optimal treatment for each individual patient. Here we describe the development of a new decisions support tool (DST) for acute appendicitis and place it in the context of international guidelines for decision aid development.

**Methods:** The stakeholder engagement and development process for the DST is described. The DST and its development process are placed in the context of the International Patient Decision Aid Standards (IPDAS) and the DEVELOPTOOLS checklist for a user-centered design process.

**Results:** A diverse group of over 60 stakeholders were involved in the needs-assessment, development, and evaluation of the DST. The development process met 11/11 of the scored items on the DEVELOPTOOLS checklist. Of the 34 applicable IPDAS items, the current version of the DST meets 31 of them including 6/6 qualifying criteria, 6/6 certification criteria, and 18/22 quality criteria.

**Conclusions:** The novel appendicitis DST was developed with the input of multiple stakeholders. The development process and the tool itself complies with best practices recommended by the IPDAS.

## Introduction

Appendicitis is one of the most common acute surgical conditions in the United States and has been largely treated with appendectomy. However, accumulating data from 9 randomized controlled trials has indicated that treatment with antibiotics with surgery if there is progression of symptoms is a safe option that will allow many patients to avoid an urgent surgical procedure.(1–9) Furthermore, data from the recent Comparison of Drugs and Antibiotic Medicines for the Treatment of Acute Appendicitis (CODA) trial(1) has shown that up to 7 in 10 patients may avoid an urgent operation and patient reported quality of life measures are similar at 30 and 90 days for participants in both treatment arms. However, there are important differences with respect to healthcare utilization, days of work missed, and risk of various complications that may influence patient decision-making and surgeon willingness to discuss and offer both treatment options.

The decision of which treatment to pursue, surgery or antibiotic therapy for acute appendicitis, requires careful consideration of multiple data points across different outcome domains in the context of the unique preferences and values of individual patients. This type of individualized patient-centered decision making may be particularly difficult for acute conditions in the emergency department given both the time-pressured nature of the decision, pain and anxiety experienced by the patient, and the fact that clinicians and patients have no prior established relationship. Furthermore, the priorities of patients and surgeons may be discordant. For example, surgeons may be most concerned about recurrence whereas patients may be most concerned about surgical pain or missing work.(10) Based on the feedback of multiple stakeholder groups involved in the CODA trial, we set out to develop a decision support tool (DST) (appyornot.org) to inform patients about their treatment options and assist patients in the emergency setting making this treatment decision. This article will describe the development process of the tool and place it specifically within the published framework of the recently published DEVELOPTOOLS checklist.(11) We also outline how the tool complies with the International Patient Decision Aid Standards (IPDAS) guideliens for a patient decision aid.(12,13)

### Intended Users and Setting

The intended users of this decision support tool are adult patients with imaging-confirmed acute appendicitis and their caregivers and the clinicians who care for them. The typical setting for use would be in the emergency department on an electronic device (either patient-owened or hospital-provided) in collaboration with the surgical consultation. Notable features of this setting include: 1) patients will typically be experiencing acute abdominal pain and may also be experiencing nausea, vomiting, or other distressing symptoms; 2) patients will typically not have a long-standing relationship with the clinicians they interact with and will often be meeting them for the first time during this encounter; 3) there is a time-pressured nature to the environment, both because appendicitis is an acute condition that requires an urgent treatment decision (that cannot be delayed and discussed at a second appointment for example) and because the emergency department often cares for a high-volume of patients and must focus on patient flow into and out of the department.

### Purpose of the Tool and Needs Assessment

Over the past number of years there has been increasing calls for enhancing the use of SDM around surgical procedures.(14,15) While shared decision making (SDM) has been embraced in many fields of surgery, emergency general surgery has for a long time been seen as a particularly difficult environment in which to implement it given that patients are often in acute distress and decisions are made in a time-presured environment. The mounting evidence for an antibiotics-first approach to treating appendicitis, has resulted in profound interest in methods to facilitate strong shared decision making for this disease process.(14) While there have been efforts to build better tools to facilitate these discussions for pediatric patients,(16,17) there has not been any tools developed to facilitate the integration of adult clinical trial data into shared decision making. This is particularly important, as the mounting evidence for antibiotic treatments may meet criteria under the Affordable Care Act requiring shared decision making.(14,15) While the most recent data from the CODA trial has shown similar outcomes for surgery and antibiotics for overall quality of life and time to symptom resolution, the treatments do differ on a number of important outcomes that may be important to patients. These outcome dimenions include days of missed work, risk of needing future surgery, and risk of missing an occult malignancy.(1) Eary discussions with patient and clinician stakeholder groups acknowledged the complexity these results may pose for effective SDM and identified the need to develop tools and procedures to facilitate it. Stakeholders recognized the variation in patient experiences and practice environments across the country and world, and the inherent variability in treatment discussions that this may lead to. They also recognized that patients and surgeons may value certain outcomes and define treatment success differently.(10) They identified key attributes of any appendicitis DST as including: effective tranmssion of knowledge in both video and text formats, facilitation of outcomes comparison between treatments, exercises to clarify the needs and values of individual patients and how they map to the different treatments and their outcomes, and finally the DST must be useable under the constraints of the typical emergency general surgery environment described above.

### Main Stakeholder Representation Groups

Figure 1 shows the main groups involved in the development of the decision support tool. The CODA trial included key stakeholders through the research cycle from conception to dissemination(18,19): a Patient Advisory Board (6 patient advisors), a Clinical Advisory Group made up of 43 surgeons and emergency medicine physicians, and a National Stakeholder Advisory Group representing professional societies, payors, and policymakers (8 members). The detailed makeup of these groups and their formation has been described previously.(19) All three of these groups served as key sources of opinions and feedback throughout the development process.

**Figure 1:**
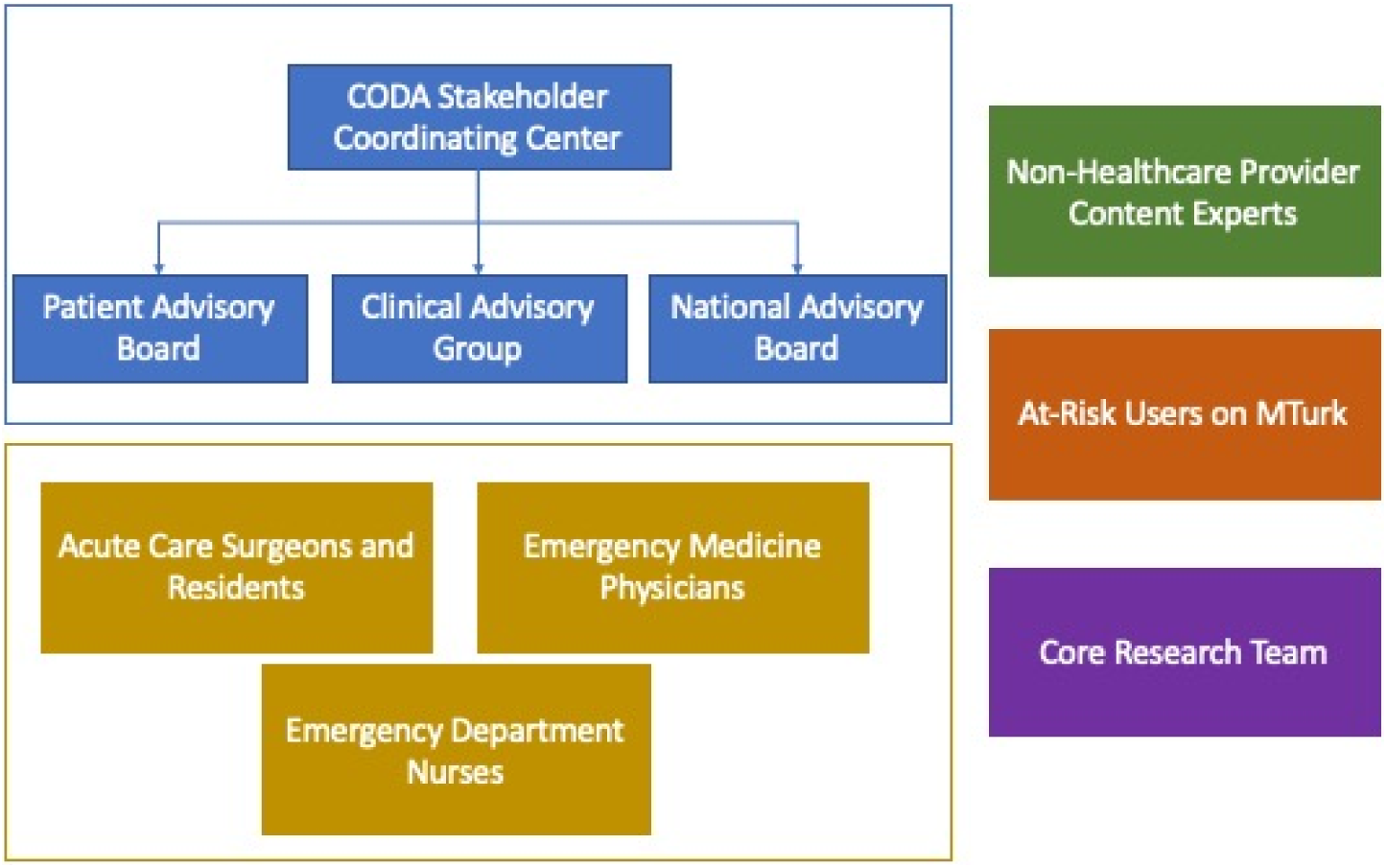
Main groups involved in the development of the appendicitis DST

**Figure 2:**
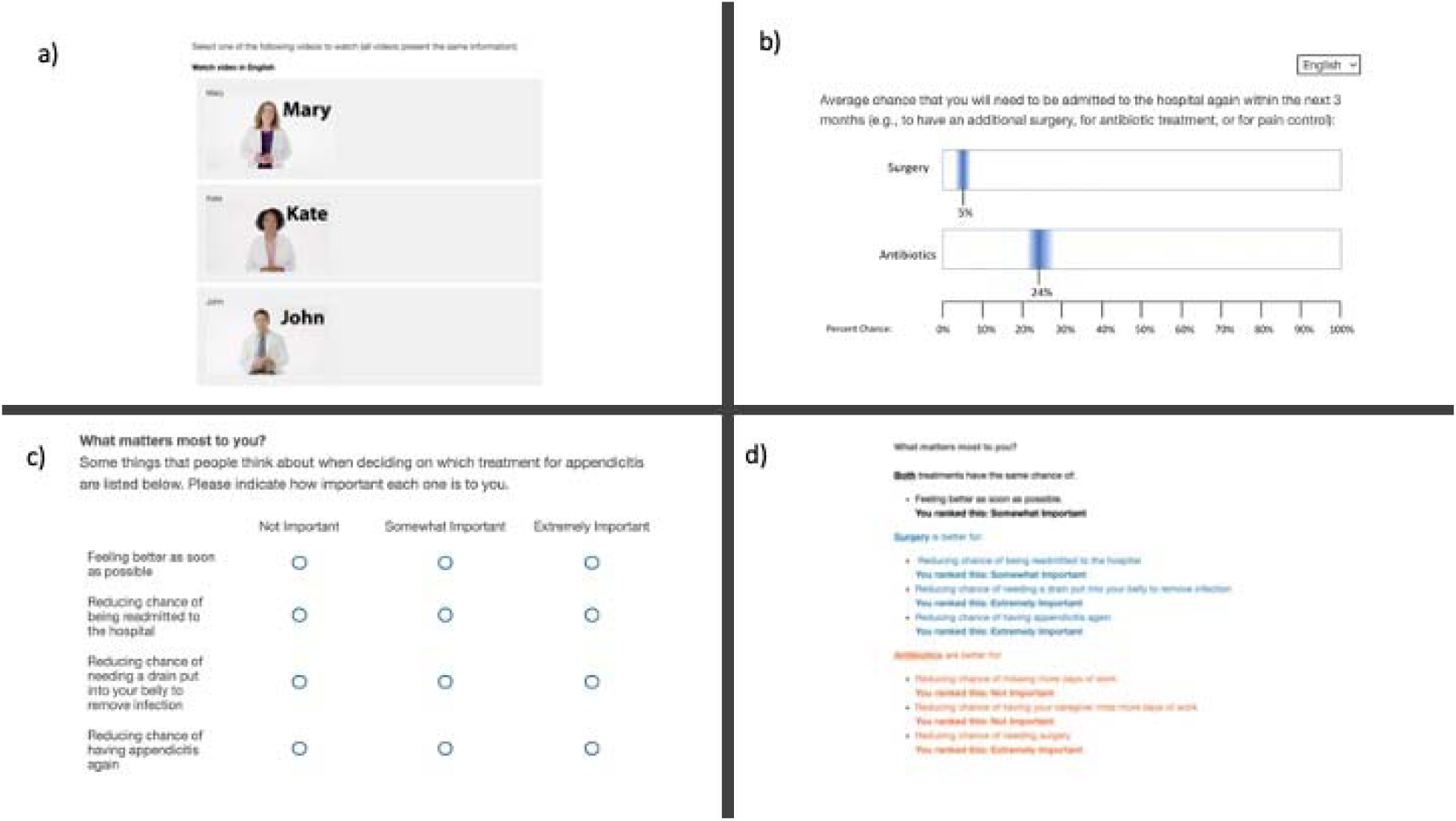
Example screenshots from decision aid a) video selection screen where users can select the informational video they would like to watch; b) data-presentation screen demonstrating the graphical format of data presentation involving point estimates and graphical confidence intervals; c) users can input what outcomes are most important to them; d) the tool categorizes those outcomes as favoring either antibiotics or surgery to aid in patient decision making.

In addition to the main stakeholder groups involved in the CODA trial, we employed several other groups in the development of this decision aid. We used online pilot-testing in people at-risk for appendicitis (i.e., no prior appendicitis, typical age distribution from the CODA trial) using Amazon’s Mechnical Turk (MTurk) crowdsourcing platform.(10,20) We engaged with non-healthcare provider content experts at our institution in the fields of cognitive and behavioral psychology, behavioral economics, and marketing to design both the decision aid elements and the testing framework used in our online pilot tests (4 people overall). We solicited opinions from a diverse group of clinical providers including acute care, surgery residents, emergency medicine physicians, and emergency department nurses. Finally, our core research team developing the decision aid included acute care surgeons, surgery residents, and experts in behavioral economics and educational video development.

### Development Process

The appendicitis DST was developed through an iterative process involving representatives of the the stakeholder groups. During analysis of the trial results,(1) multiple focus groups were held with patient and physician stakeholders focusing on the interpretation of the trial results and planning for dissemination and implementation strategies. Stakeholders provided input on the purpose and goals of an appendicitis DST and the concept of a DST consisting of an informational video followed by an interactive decision aid was solidified. Furthermore, CODA stakeholder groups identified the key outcome data points for inclusion in the decision aid and video.

The informational video was developed based on a prior video that had been created to facilitate a more uniform informed consent process for participation in the CODA trial.(21) The video script underwent multiple rounds of development and review by patient and clinician takeholders, as well as review by healthcare communications experts. Finally, the videos were professionally shot and edited and final videos were reviewed by key stakeholders prior to inclusion in the DST.

The interactive patient decision aid was developed by using the needs assessment specified by the CODA advisory groups to complete a literature search identifying best practices in the key areas of information communication and values clarification. We also consulted with non-clinicina experts as described above. We then performed five rounds of iterative and sequential testing of our information delivery practices using over 1,300 subjects on MTurk. After developing an initial prototype based on this iterative testing, we solicited feedback from CODA stakeholder groups, and the additional clinical stakeholders identified in Figure 1.

### Description of Decision Aid

The DST consists of two main components, an informational video, and an interactive decision aid. Patients first view a professionally produced video in either English or Spanish that describes the pathophysiology of appendicitis, reviews evidence-based treatment options (e.g., antibiotics and surgery), and reviews information on the expected clinical course. Following the video, patients view the interactive decision aid which reviews clinical outcomes comparing antibiotics and surgery followed by a values-clarification exercise where patients rate the importance of various outcomes and see how they align with each treatment option. Finally, a summary screen is provided that can be used to help facilitate discussion of treatment options with the clinical team and patient.

### Assessing the Development Process – The DEVELOPTOOLS Checklist

The IPDAS group provides a basis for a recommended systematic development process and a recently published update incorporates principles of user-centered design to create the DEVELOPTOOLS checklist(11) for reporting the design and development process for patient decision aids. In Table 1 we outline the components of the development process for the appendicitis DST using the DEVELOPTOOLS checklist.

**Table 1:** DEVELOPTOOLS checklist and evidence items from the Appendicitis DST

| **<u>Checklist Item</u>** | **<u>Evidence from Appendicitis DST</u>** | **<u>Scoring</u>**<br>**<u>(1 =</u>**<br>**<u>Present,</u>**<br>**<u>0 =</u>**<br>**<u>Absent)</u>** |
| --- | --- | --- |
| **Pre-prototype Involvement** |  |  |
| Were potential users involved in any steps to help understand users and their needs? | Yes, members of the CODA patient advisory board, clinical advisory group and national stakeholder advisory group (see above for numbers) were involved. emergency department nurses and surgery residents were involved. | 1 |
| Were potential users involved in any steps of designing, developing, and/or refining a prototype? | Yes, see Figure 1 and Stakeholder section above for all groups involved in prototype development. | 1 |
| **Iterative Responsiveness** |  |  |
| Were potential users involved in any steps to evaluate prototypes of the tool or a final version of the tool? | Yes. We used online crowd-sourcing via MTurk (over 1200 subjects), as well as the clinical and patient advisors described in Figure 1. Furthermore, we are actively trialing the pilot version of the tool with patients in the emergency department at our institution. | 1 |
| Were potential users asked their opinions of prototypes of the tool or a final version of the tool in any way? | Yes, see Figure 1 and stakeholder section above. | 1 |
| Were potential users observed using the tool in any way? | Yes, surgeons and patients are currently being observed using the tool in the emergency department. | 1 |
| Did the development process have 3 or more iterative cycles? (Definition of a cycle is that your team developed something and showed it to at least one person outside of the team before making changes in response to their feedback) | Yes, there was 5 cycles of online development studies and at least 3 more cycles involving feedback from the other clinician and patient stakeholder in Figure 1. | 1 |
| Were changes between iterative cycles explicitly reported in any way? | Yes in internal logs | 1 |
| **Other Expert Involvement** |  |  |
| Were health professionals asked their opinion of the tool at any point? | Yes the clinical advisory group as well as additional acute care surgery faculty, surgery residents, and emergency medicine physicians were asked about the tool. | 1 |
| Were health professionals consulted before a first prototype was developed? | Yes, as described above the clinical advisory group was consulted prior to prototype development. | 1 |
| Were health professionals consulted between initial and final prototypes? | Yes the clinical groups in Figure 1 were consulted at multiple points. | 1 |
| Was an expert panel involved? | Yes, as described in the stakeholder section above. | 1 |
| **Additional Elements** |  |  |
| Was a formal advisory panel of users involved? | Yes, see(19) for information on the assembly of stakeholder panels. | N/A |
| Were users, health professionals, and other relevant stakeholders involved as members of the research team? | Yes, acute care surgeons and marketing/behavioral economics experts were research team members. Furthermore, the CODA patient advisors are considered to be members of the research team and contributed to the results dissemination discussions. | N/A |
| Were members of populations marginalized by social norms and policies involved? | Unable to answer, under-represented minorities were under-recruited to the patient advisory board. | N/A |
| How many users and health professionals were involved in total and of each type? | See stakeholder section and Figure 1 above. | N/A |
| Does the tool have a defined purpose? | Yes, the tool's purpose is to provide consistent information about treatment options for appendicitis and to help patients decide between surgical and | N/A |
|  | antibiotic treatment |  |
| Is the tool intended to be used in a particular context? | Yes, the tool is intended to be used in the emergency setting. | N/A |
| Were any methods used to facilitate sharing of perspectives between groups? | Yes, stakeholder meetings often were cross-disciplinary and both structured and open ended questions were asked of participants. Deidentified summary of discussion points were also shared. | N/A |
| Were users involved from the outset of the project? | Yes, patient and clinician users have been involved from the outset of the CODA project | N/A |
| Were translations and cultural adaptation used to render the patient decision aid available to users across languages and cultures? | Yes, for the videos we have multiple actors from different cultural backgrounds as well as a Spanish language option. The decision aid is translated into Spanish. Additional languages including American Sign Language are planned. | N/A |

### Assessment of Decision Support Tool by IPDAS Guidelines

The IPDAS provides a framework for the evaluation of patient decision aids. Version 4.0 published in 2013 specifies a set of minimum criteria broken up into three categories: qualifying criteria (defining if a tool is a decision aid), certification criteria (defining essential facets of a decision aid), and quality criteria (those that will enhance a decision aid but are not essential).(12) Table 2 below shows the 34 applicable criteria in IPDAS 4.0 (9 of the standard 44 criteria are for decision aids involving the use and interpretation of medical tests, 1 additional criterion is not applicable to the appendicitis DST) and whether or not the appendicitis DST meets them in its current form. Overall 6/6 qualifying criteria are met, 6/6 certification criteria are met, and 18/22 quality criteria are met, with three currently in progress.

**Table 2:**
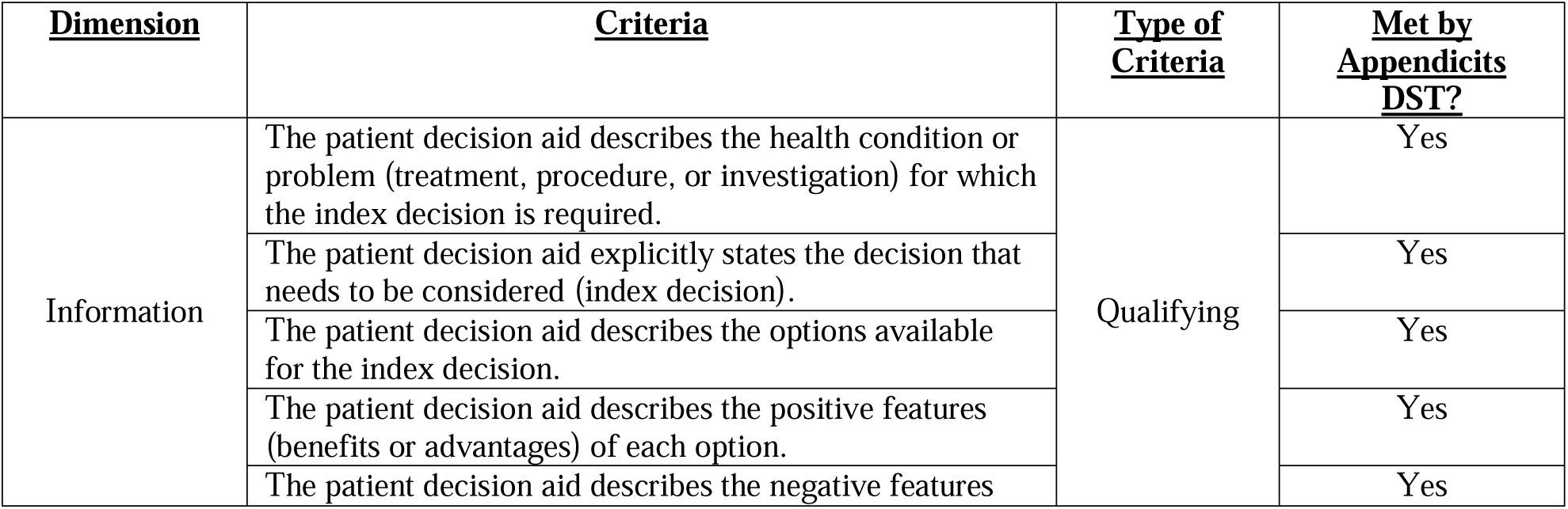
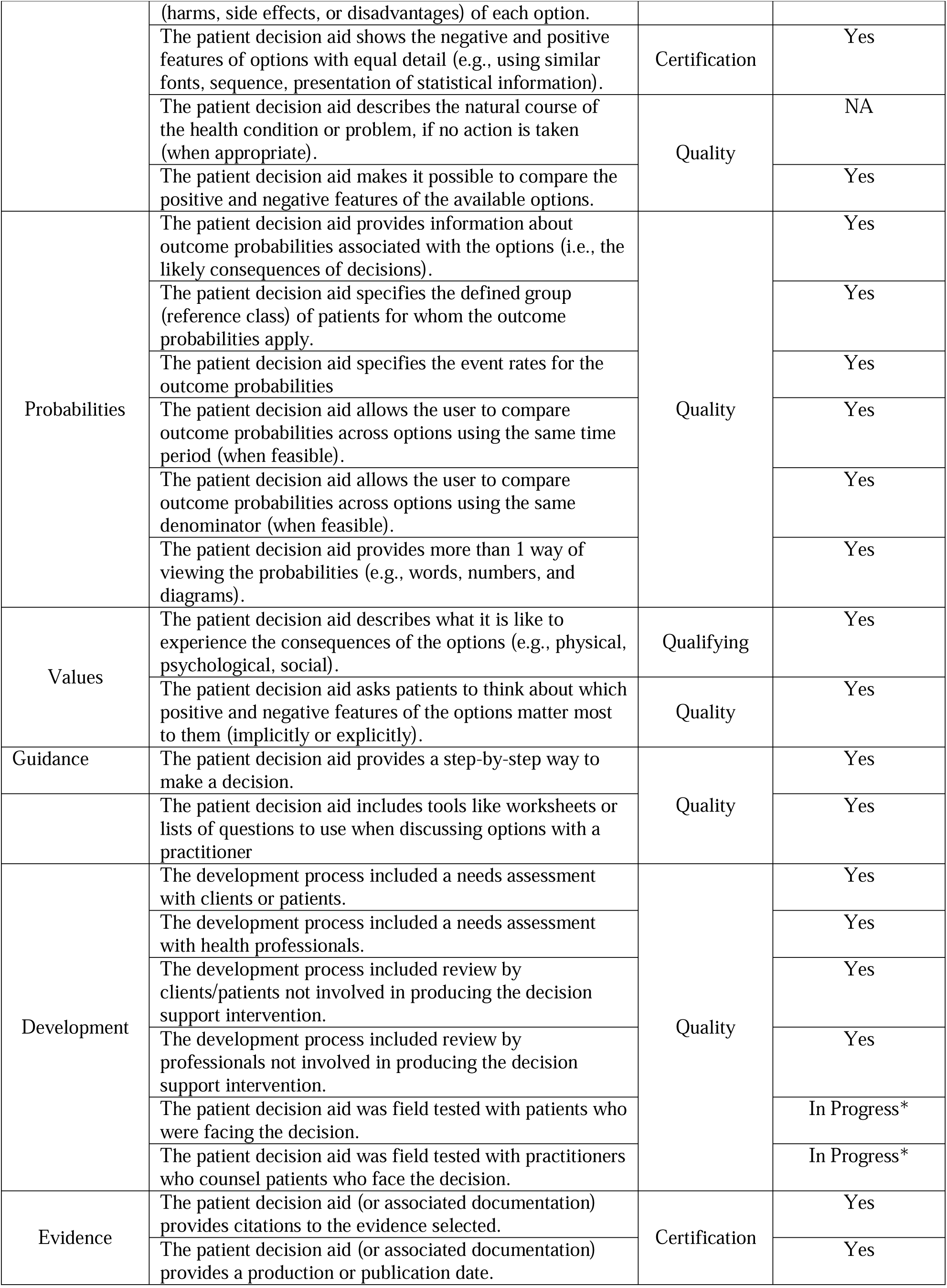
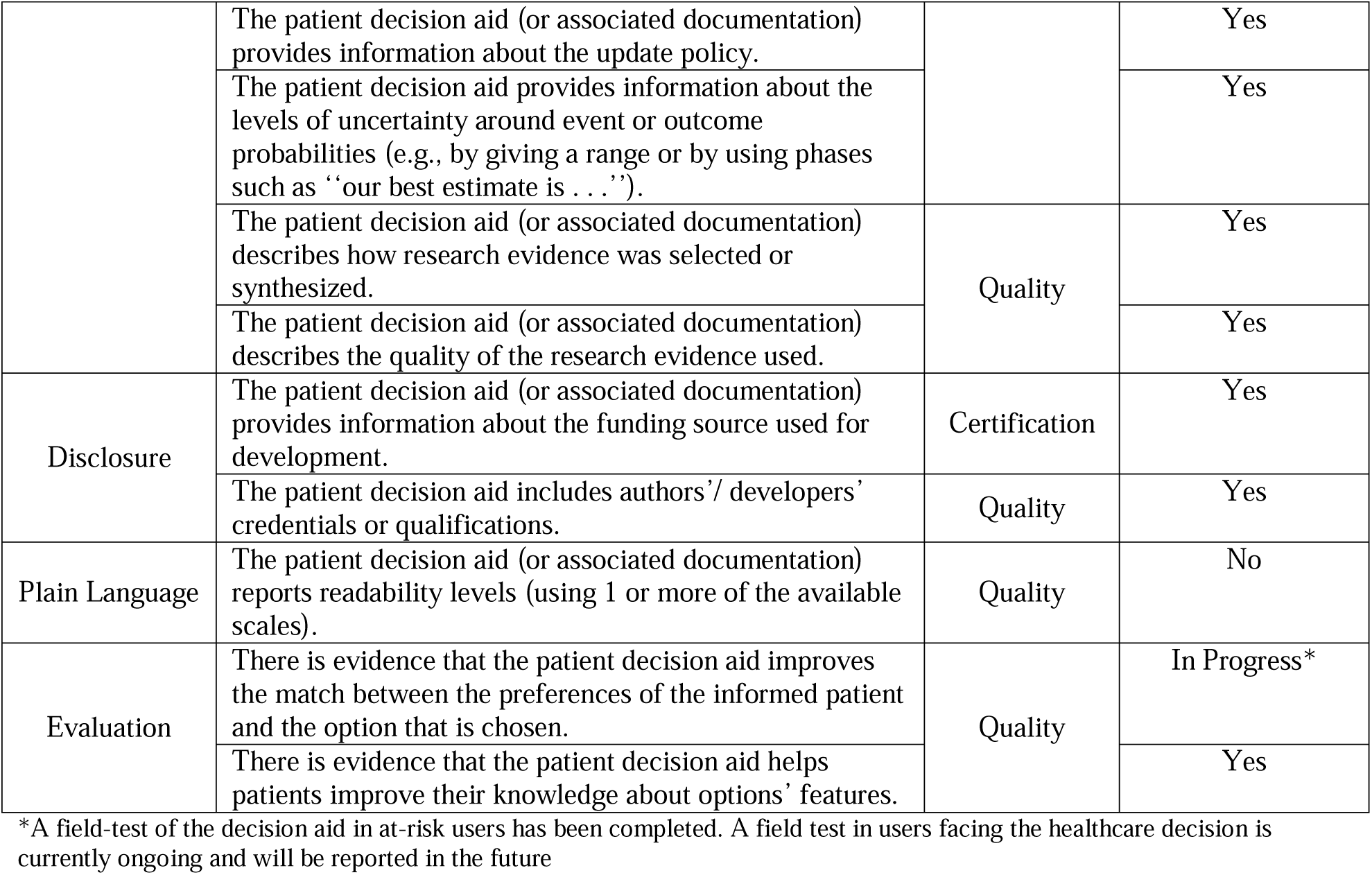
IPDAS 4.0 criteria and scoring of appendicitis DST

## Conclusions

In this article we have described the development process for a new decision aid for acute appendicitis. The process involved input from multiple clinician and patient stakeholders and adhered to principles of user-centered and evidence-based design as described in the DEVELOPTOOLS checklist. The resulting decision support tool complies with the IPDAS 4.0 criteria. To our knowledge this is one of the first DSTs specifically developed for use in adult emergency general surgery patients. This may serve as a roadmap to more efficiently develop patient-centered tools for SDM for emergency surgical patients.

## Data Availability

All data produced in the present study are available upon reasonable request to the authors

